# Protocol for Safe, Affordable, and Reproducible Isolation and Quantitation of SARS-CoV-2 RNA from Wastewater

**DOI:** 10.1101/2021.02.16.21251787

**Authors:** Monica Trujillo, Kristen Cheung, Anna Gao, Irene Hoxie, Sherin Kannoly, Nanami Kubota, Kaung Myat San, Davida S. Smyth, John J. Dennehy

**Affiliations:** Department of Biology, Queensborough Community College, The City University of New York, New York; Biology Department, Queens College, The City University of New York, New York; The Graduate Center, The City University of New York, New York; Department of Natural Sciences and Mathematics, Eugene Lang College of Liberal Arts at The New School, New York.

**Author notes:** These authors are listed in alphabetical order (Kristen Cheung, Anna Gao, Irene Hoxie, Sherin Kannoly, Nanami Kubota, Kaung Myat San, Davida S. Smyth).

## Abstract

The following protocol describes our workflow for processing wastewater with the goal of detecting the genetic signal of SARS-CoV-2. The steps include pasteurization, virus concentration, RNA extraction, and quantification by RT-qPCR. We include auxiliary steps that provide new users with tools and strategies that will help troubleshoot key steps in the process. This protocol is one of the safest, cheapest, and most reproducible approaches for the detection of SARS-CoV-2 RNA in wastewater. Furthermore, the RNA obtained using this protocol, minus the pasteurization step, can be sequenced both using a targeted approach sequencing specific regions or the whole genome. The protocol was adopted by the New York City Department of Environmental Protection in August 2020 to support their efforts in monitoring SARS-CoV-2 prevalence in wastewater in all five boroughs of the city. Owing to a pasteurization step, it is safe for use in a BSL1+ facility. This step also increases the genetic signal of the virus while making the protocol safe for the personnel involved. This protocol could be used to isolate a variety of other clinically relevant viruses from wastewater and serve as a foundation of a wastewater surveillance strategy for monitoring community spread of known and emerging viral pathogens.

## Introduction

The tracking of SARS-CoV-2 infections has most often involved the detection of SARS-CoV-2 RNA via RT-qPCR in biological samples obtained from patients that develop some of the symptoms associated to COVID-19 [1]. One of the disadvantages of this approach is that if much of the transmission within a population is asymptomatic or unsampled, infections from these individuals may be overlooked [2,3]. Additionally, SARS-CoV-2 sequencing efforts, while occurring at a much faster rate and larger, more global scale than in previous pandemics, suffer biases because genomic information is often obtained from seriously ill patients, but not from patients who do not seek medical attention, which include asymptomatic patients, and those with mild symptoms who choose to follow the CDC’s advice and convalesce at home. If most transmission within a population is asymptomatic or unsampled, genomes from these individuals are expected to represent most of the viral population circulating within the community. Recently the discovery of novel variants of concern in different regions of the world has added another challenge [4,5], which is to monitor the proportion of individuals that carry a particular variant in a geographical area. Given that SARS-CoV-2 has been detected in fecal samples [6,7], and subsequently in wastewater [3,8,9], wastewater is being tested in cities around the world to determine SARS-CoV-2 prevalence in communities [10–12]. Furthermore, isolation of SARS-CoV-2 RNA from wastewater coupled with high-throughput deep sequencing provides an almost unlimited source of unbiased viral sequences, which can be used to monitor frequencies of variants of concern in populations.

With the goal of sequencing SARS-CoV-2 RNA from wastewater, we developed a protocol to extract and quantify viral RNA. The initial step in the development of this protocol was the decision to pasteurize our samples at 60 □ for an hour on arrival at the laboratory. Given that SARS-CoV-2 is a biosafety level 3 (BSL3) agent, inactivation of the virus before processing is often required before samples can be processed in BSL2+ or BSL1+ laboratories. Happily, as we report here, pasteurization did not impair our ability to detect SARS-CoV-2, but instead, improved it. Interestingly, while SARS-CoV-2 recovery was not impaired, control spike-in viruses bovine coronavirus (BcoV) [13] and bacteriophage Phi6 [14] were barely detectable using RT-qPCR and PCR respectively. Subsequently, control viruses were spiked-in after pasteurization. We are currently studying the effect of pasteurization on the quality of our sequencing data. Preliminary results suggest that the output and quality of sequencing data may be better with unpasteurized samples, therefore if the intention of the study is to sequence SARS-CoV-2 from wastewater, we recommend skipping the pasteurization step.

A second major decision was to employ centrifugation and filtering (0.2 µM) to remove the wastewater solids. While it was acknowledged that SARS-CoV-2 may associate with the solids, removing the solids facilitates downstream processing steps, and may remove genomic contamination that would impair our ability to deep sequence SARS-CoV-2. As a counterpoint, filtration is one of the more expensive steps of the protocol so those desiring to reduce costs may consider eliminating filtration. We were able to acquire consistent results with and without filtration, and neither strategy resulted in a significant increase in our ability to quantify SARS-CoV-2.

Since viruses are greatly diluted in wastewater, virion concentration is a significant challenge. We considered three common protocols to concentrate SARS-CoV-2 virus present in the water: ultracentrifugation [15], skimmed milk flocculation [16], and polyethylene glycol (PEG)/sodium chloride (NaCl) precipitation. High speed centrifugation was ruled out as impractical for the volumes needing to be processed. Precipitation/flocculation using PEG/NaCl or skimmed milk eliminates the need for high-speed ultracentrifugation and generates sufficient RNA for viral quantification with RT-qPCR (i.e., resulting in Cts < 40). However, in our experiments, PEG/NaCl precipitation performed marginally better than skim milk flocculation and does not introduce additional genetic material to our samples, so this was chosen as our concentration method. As we expanded our experiment to include sequencing the RNA from wastewater, we explored the effect of longer incubation times on viral RNA recovery. Longer storage in PEG/NaCl led to slightly greater recovery.

As we were mindful of the need to find cost effective solutions, we investigated alternative, kit-free approaches to RNA isolation. In our hands, TRIzol (ThermoFisher Inc.) performed better than the QIAamp Viral RNA Mini kit (Qiagen Inc.). As TRIzol is cheaper per sample than column-based kits, we adopted it for the final protocol. An added benefit of TRIzol relevant to downstream sequencing applications is that TRIzol segregates RNA in a separate layer from DNA, unlike column-based isolation kits, which isolate both RNA and DNA.

In addition to the RNA isolation method, we compared the performance of different RT-qPCR enzymes, TaqPath 1-Step RT-qPCR enzyme (Thermofisher Inc.) and One Step PrimeScript III enzyme (Takara Bio USA Inc. The RT enzyme from Takara was 25% cheaper and had a similar performance to Taq-Path so we chose it for the final protocol. A broader investigation of different enzymes may identify other satisfactory, cost-effective solutions.

Our protocol provides a reproducible and low-tech approach that allows the detection and quantification of SARS-CoV-2. Pasteurization of the sample at the very beginning of the protocol ensures the safety of the user. Preliminary results suggest that pasteurization may also release the virus bound to the wastewater solids, enhancing recovery. Filtering and PEG/NaCl concentration simplifies downstream processing. The extraction of RNA using TRIzol reduces the cost significantly when compared to extraction column-style protocols using commercial kits. We have been able to do both targeted and whole genome sequencing of the SARS-CoV-2 genome using this protocol but recommend removing the pasteurization step if this is the main goal of the experiments.

Our protocol performed strongly in a large-scale, nationwide comparative study of the reproducibility and sensitivity of 36 methods of quantifying SARS-CoV-2 in wastewater [17]. Our protocol is identified as 4S.1(H) in Table 3. In addition, the Pecson et al. study offers strong support for several of the primary claims of the present paper. First, the removal or non-removal of the wastewater solids did not show a clear systematic impact on outcomes. Second, pasteurization resulted in a small, but significant, increase in recovery. Third, methodological differences between teams had minimal impact on reproducibility and sensitivity, thus indicating that our modifications to implement cheaper, simpler methods will not impair SARS-2-CoV-2 detection and quantification relative to other strategies.

We recognize that our protocol has some limitations. Our current protocol isolates the RNA from 40 ml of wastewater and requires access to a centrifuge capable of reaching 12,000 x g. Thus, scaling up the volume of samples from 40 ml or increasing the number of individual samples, represents a challenge. Our protocol requires filtration units which are dependent on the supply chain. Additionally, extracting RNA with TRIzol requires the user to take care not to contaminate the aqueous phase with organic material after centrifugation, which can be difficult for inexperienced users. Nevertheless, the basic protocol and techniques involved is economical, simple, and reproducible when compared to alternative strategies.

## Materials and Methods

*“The protocol described in this article is published on protocols.io, dx.doi.org/10.17504/protocols.io.brr6m59e and is included for printing as file S1*.*”*

### Expected Results

Our protocol results in the reproducible isolation and quantification of SARS-CoV-2 RNA from wastewater samples (Fig. 1). Enough RNA can be acquired for RT-qPCR, and isolated RNA is suitable for whole genome amplification and sequencing (although pasteurization is not recommended if the intention is to sequence SARS-CoV-2 RNA isolated from wastewater). As a general note, wastewater treatment plants indicated in our figures have been deidentified. There is no correspondence between the numerical wastewater treatment plant (WWTP) IDs in different figures. Moreover, experiments described in different figures were performed at different times using different wastewater samples. Our purpose here is not to report regional prevalence, but rather to demonstrate the reliability and consistency of our protocol.

**Figure 1.**
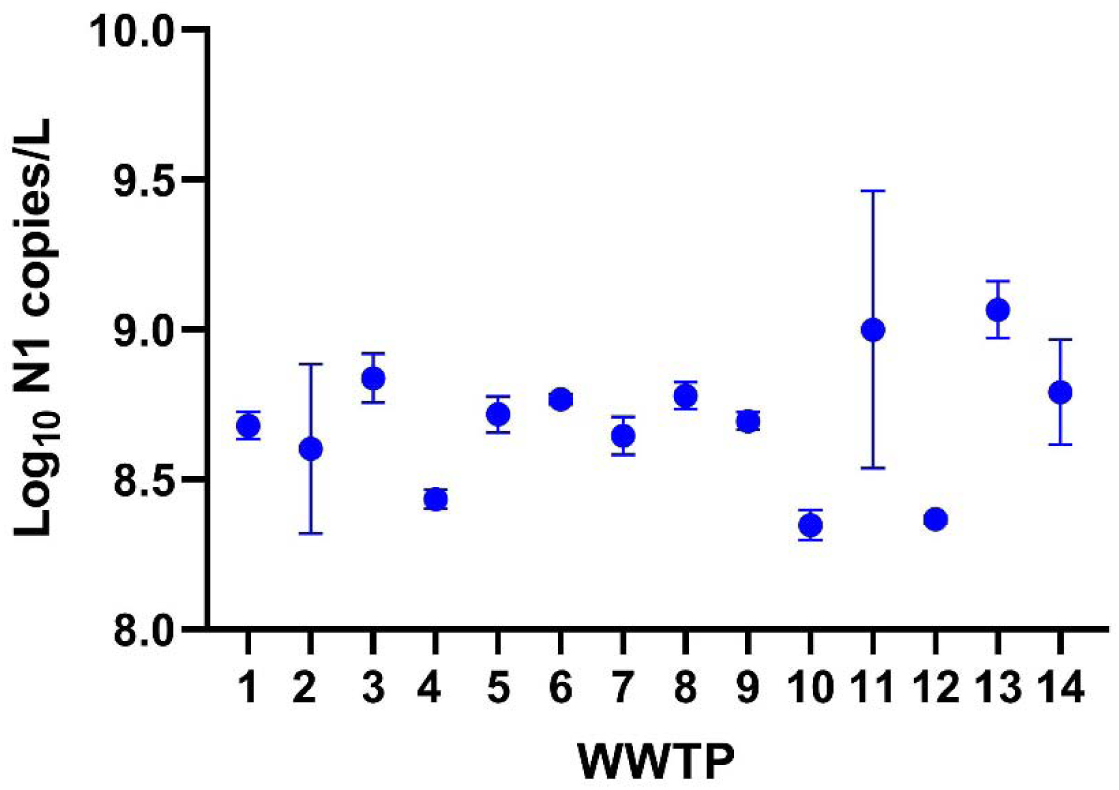
Repeatability of Protocol: Copy number yield for the N1 target for 24-hr composite wastewater samples obtained from all 14 wastewater treatment plants (WWTP) in New York City demonstrating reproducibility of our protocol. Each point is the mean of two technical replicate measurements from a 24-hour composite sample. All samples collected and initially processed on the same day. Error bars are ±SEM. Some error bars are too small to be visible. Samples 2 and 11 are from plants with a significant influx of ocean water, but it is not clear if this is driving variation in these sites.

Key steps were optimized during the development phase of our protocol. Initially we used Phi6 [14] as a spike in control. However, we found that Phi6 was rapidly degraded in the pasteurization step of our protocol. Reports from the scientific community suggested that BCoV would serve as a better control, however, we found that BCoV was significantly degraded by pasteurization as well. Consequently, we switched to spiking samples with BCoV after pasteurization and before the first centrifugation to remove solids. It would be interesting to determine why BCoV was rapidly degraded by pasteurization, but an ostensibly similar virus, SARS-CoV-2, was not.

To ascertain the impact of pasteurization on SARS-CoV-2 quantitation, a single California wastewater sample was divided into ten parts. Five of these replicate samples were pasteurized and five were not. The positive impact of pasteurization on SARS-CoV-2 quantification is reflected in the increase of N1 copies/L of pasteurized versus unpasteurized samples (Fig. 2; paired t-test: t = 7.191, df = 4, p = 0.002). We speculate that incubation of samples at 60 LJ contributes to release of virus from wastewater solids. As an additional advantage, pasteurization appears to increase repeatability of sample quantification. The pooled variance for pasteurized and unpasteurized samples were 0.005 and 0.177 respectively. We conclude that pasteurization results in greater sensitivity and more precise estimates of SARS-CoV-2 prevalence. Similar outcomes have been reported elsewhere [17].

**Figure 2.**
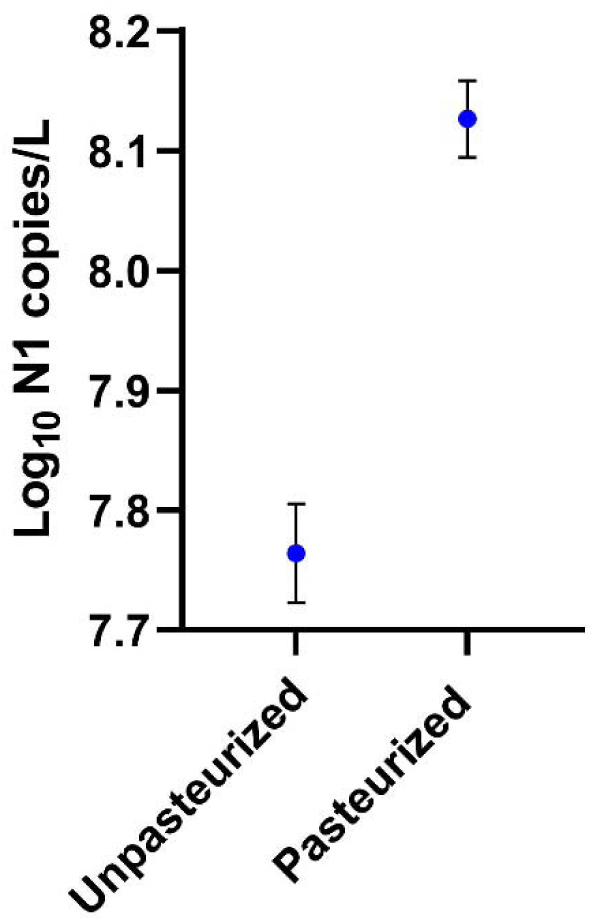
Effect of Pasteurization: Copy number yield for the N1 target obtained from one California 24-hr composite wastewater sample processed either with pasteurization or without pasteurization. Each point is the mean of 5 independent assays. Error bars are ±SEM. A paired t-test revealed significant differences between the treatments (t = 7.191, df = 4, p = 0.002).

In previous work on bacteriophages, we had observed that longer PEG/NaCl incubation times increased phage recovery. To determine if longer incubation similarly impacts SARS-CoV-2 recovery, we compared SARS-CoV-2 quantitation for samples incubated in PEG/NaCl for 24 hrs versus 48 hrs. We found that 48 hrs incubation significantly increased sample yield (Fig. 3; RM ANOVA: F = 398, P = 0.0003). This result should ameliorate concerns about longer term storage of wastewater samples if they cannot be processed immediately. In our hands, SARS-CoV-2 quantitation was not impaired in the short term by storage at 4 °C either with or without PEG/NaCl, an outcome similarly reflected by other studies [18, 19]. However, storing the pasteurized samples at 4 □ for 72 hours without added PEG and NaCl negatively impacted the recovery of N1 copies by RT-qPCR.

**Figure 3.**
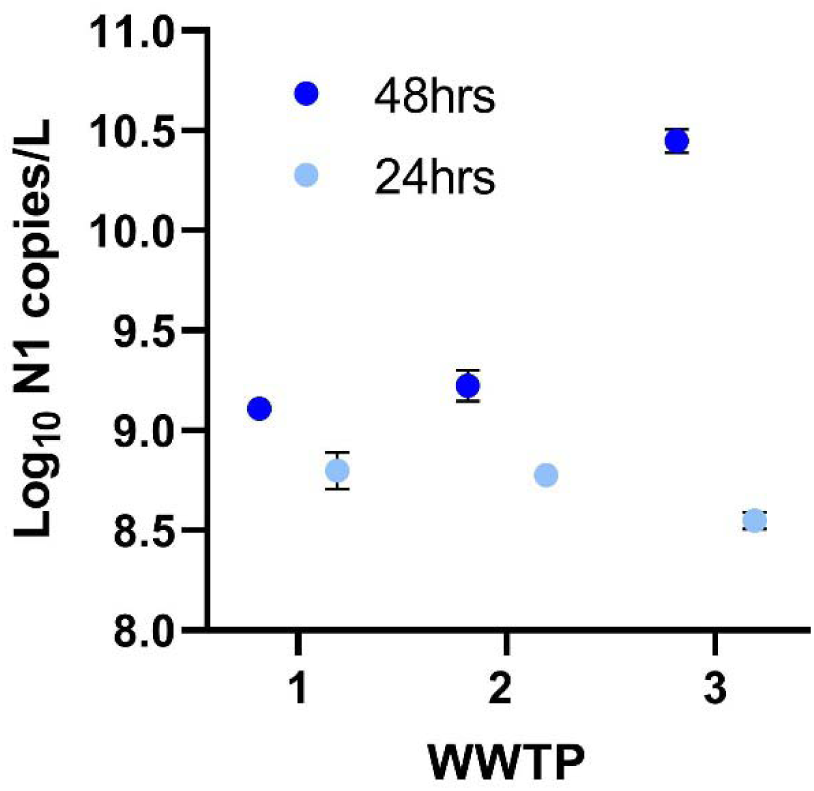
Effect of Storage Time: Following initial processing (pasteurization, preliminary centrifugation, and filtering), 24-hr composite samples from 3 different wastewater treatment plants were stored in a PEG/NaCl solution for precipitation and concentration of virions. Samples stored in PEG/NaCl solution for 48-hrs are labeled by dark blue circles; samples stored in PEG/NaCl solution for 24-hrs are labeled by light blue circles. Each point is the mean of two technical replicate measurements from a 24-hour composite sample. Error bars are ±SEM. Some error bars are too small to be visible. A repeated measures ANOVA indicated that hrs storage in PEG/NaCl resulted in significantly greater yields than did storage for 24-hrs (F = 398, P = 0.0003).

The pellet obtained after centrifugation of the wastewater sample (with added PEG and NaCl) is not visible to the naked eye in most cases and is usually distributed along the side of a polypropylene Oak Ridge tube. Additionally, it takes time to dissolve the pellet in TRIzol, and premature decanting may leave residual RNA unrecovered. Therefore, untrained users often resuspend the pellet incompletely, resulting in the loss of valuable RNA. To aid in visualizing the pellet, we added safranin at 0.2% final concentration immediately before centrifugation. Safranin did not interfere with downstream processing (Fig. 4). When safranin is added, a pale pink pellet is easily visible. The video uploaded as Supplementary Material (S2) shows how long it takes to dissolve the pellet in TRIzol. This strategy of adding safranin is particularly useful for training purposes.

**Figure 4.**
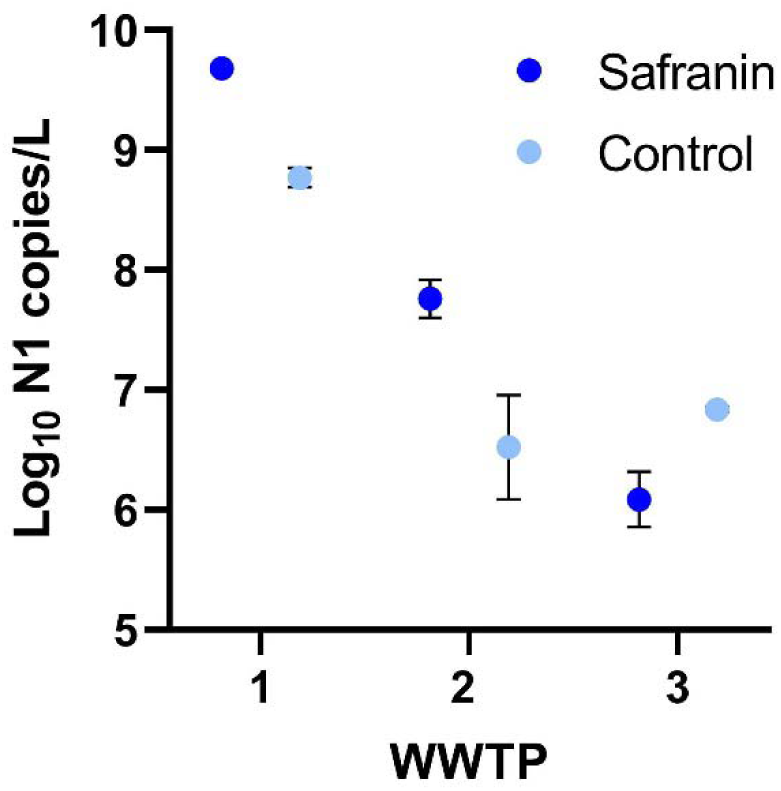
Effect of Safranin Staining: Copy number yield for the N1 target for 24-hr composite wastewater samples obtained from 3 wastewater treatment plants. Samples processed with safranin are labeled by dark blue circles; controls are labeled with light blue circles. Each point is the mean of 4 technical replicate measurements from a 24-hour composite sample. Error bars are ±SEM. Some error bars are too small to be visible. A repeated measures ANOVA revealed that safranin staining improved virus recovery (F= 15.10, P = 0.006). This effect is likely a result of better visibility of the RNA pellet during recovery.

To explore the cheapest alternatives of extracting RNA from wastewater samples we compared a widely used column-based QIAamp Viral RNA Mini kit (Qiagen Inc.) with TRIzol (ThermoFisher Inc.). TRIzol facilitates significantly better RNA recovery than the kit at a fraction of the cost (Fig. 5; RM ANOVA: F= 1441, P < 0.0001). We note that we also found phenol-chloroform extraction to be less consistent than TRIzol on saliva samples, so while phenol-chloroform is likely even cheaper, we advise against its use in this protocol. TRIzol was therefore chosen as the organic extraction method to compare with column approaches. Importantly the supply of TRIzol is less impacted by supply chain issues. Additionally, TRIzol removes DNA, but retains RNA, whereas column-based kits are unable to do so. If the intention is to sequence RNA obtained from wastewater samples, TRIzol extraction provides a cleaner sample with less contaminating DNA from non-SARS-CoV-2 genomes. As a caveat, because TRIzol requires the careful extraction of an aqueous layer from a multilayered solution, TRIzol extraction requires training and is best performed by experienced users.

**Figure 5.**
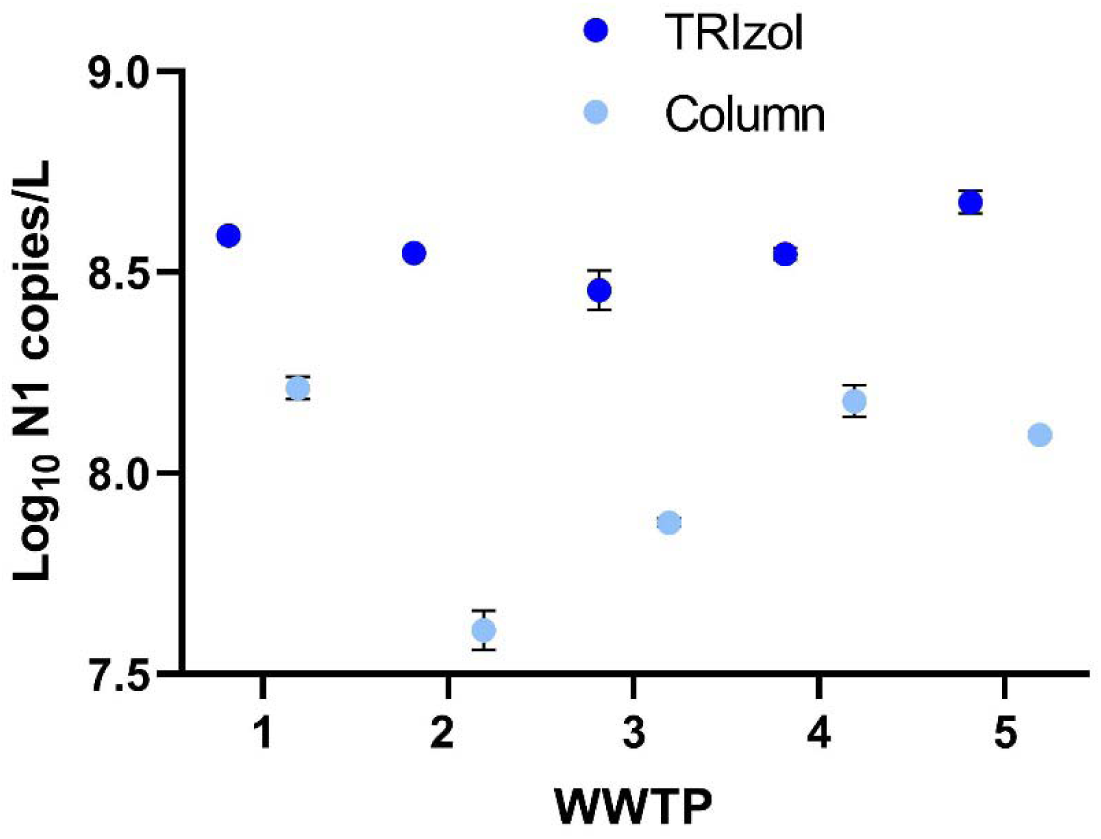
Effect of TRIzol Extraction: Copy number yield for the N1 target for 24-hr composite wastewater samples obtained from 5 wastewater treatment plants. Samples processed with TRIzol are labeled by dark blue circles; samples processed with the QIAamp Viral RNA Mini Kit are labeled with light blue circles. Each point in the mean of 2 technical replicate measurements from a 24-hour composite sample. Error bars are ±SEM. Some error bars are too small to be visible. A repeated measures ANOVA revealed that the use of TRIzol significantly improved virus RNA recovery (F= 1441, P < 0.0001).

In addition to comparing RNA isolation methods, we evaluated the performance of different enzymes, including the TaqPath 1-Step RT-qPCR enzyme (ThermoFisher Inc.) and One Step PrimeScript III enzyme (Takara Bio USA Inc.) Our results indicated that the One Step PrimeScript III enzyme gave slightly better results (Fig. 6). As the One Step PrimeScript III enzyme was 25% cheaper and performed similarly to the ThermoFisher enzyme, we chose the PrimeScript III enzyme for the final protocol.

**Figure 6.**
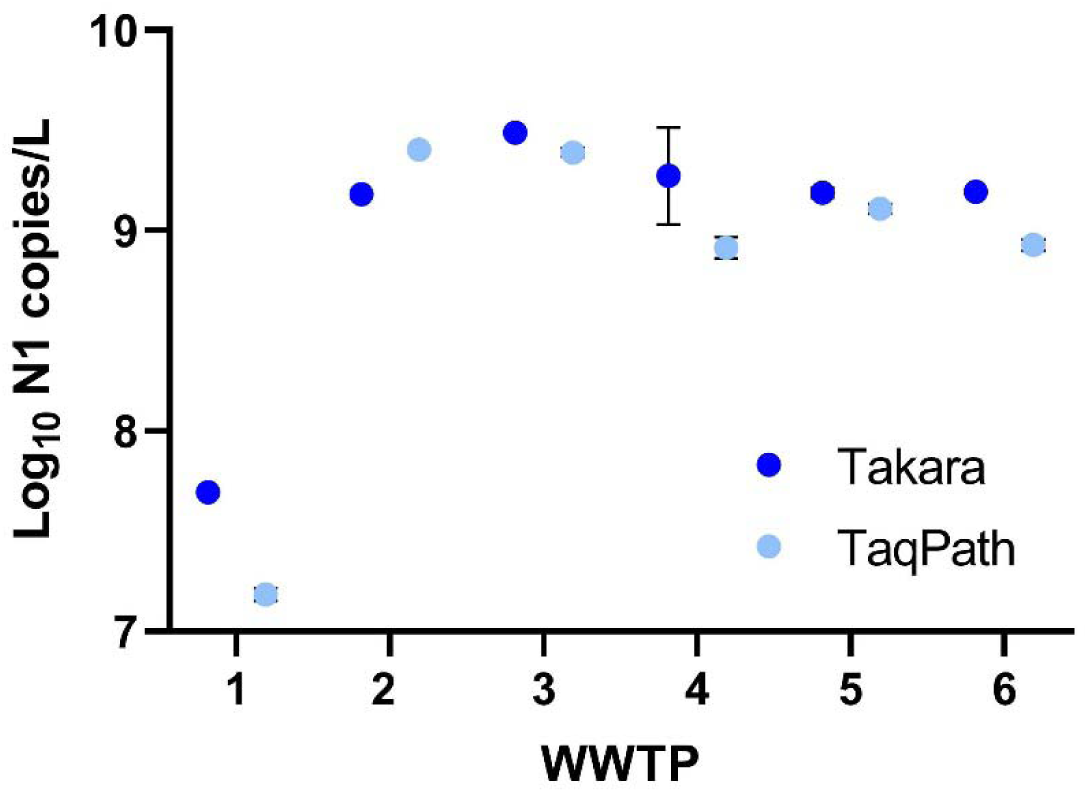
Effect of Different RT-qPCR Enzymes: Copy number yield for the N1 target for 24-hr composite wastewater samples obtained from 6 wastewater treatment plants (WWTP). RT-qPCR assays performed with the TaqPath 1-Step RT-qPCR enzyme (ThermoFisher Inc.) recommended by the CDC [20] are labeled by light blue circles; RT-qPCR assays performed with One Step PrimeScript III enzyme (Takara Bio USA Inc.) are labeled with dark blue circles. Each point in the mean of 2 technical replicate measurements from a 24-hour composite sample. Error bars are ±SEM. Some error bars are too small to be visible. A repeated measure ANOVA revealed that the use of the PrimeScript III enzyme improved virus RNA recovery (F= 13.09, P = 0.011).

The need to adapt wastewater surveillance detection programs to include variant detection requires deep sequencing of cDNA generated from the wastewater RNA. Our preliminary results have shown that RNA extracted with our PEG/TRIzol protocol can be sequenced using both traditional Sanger sequencing and NGS technology. However, we also note that pasteurization reduced sequencing quality and output relative to unpasteurized samples, so we recommend skipping this step if the intention is to sequence RNA obtained from wastewater.

We used both the Swift Normalase^®^ Amplicon Panel (SNAP) SARS-CoV-2 Panel kit as well as the Qiagen QIAseq® SARS-CoV-2 Primer Panel and QIAseq FX DNA Library kit and have obtained SARS-CoV-2 sequences from several of our wastewater treatment plants. Known and novel variants were identified. We continue to optimize and improve our library preparation methods to increase both length of coverage and depth of coverage for our NYC samples. In addition, we are developing real-time assays for the identification and quantification of additional viruses that circulate among our New York communities including Influenza.

## Supporting information

Safranin staining to visualize RNA pellet

## Data Availability

All data associated with this manuscript are available on https://datadryad.org/: https://doi.org/10.5061/dryad.zkh189396.

https://www.protocols.io/view/protocol-brr6m59e/

https://doi.org/10.5061/dryad.zkh189396

## Acknowledgements

The research described herein would not be possible if not for the assistance and support of a wide-range of organizations and individuals that came together to address the shared calamity that is the COVID-19 pandemic. We thank Gina Behnke, Esmeraldo Castro, Francoise Chauvin, Alexander Clare, Pilar Domingo-Calap, Pam Elardo, Raul Gonzalez, Crystal Hepp, Catherine Hoar, Dimitrios Katehis, William Kelly, Samantha McBride, Hope McGibbon, Timon McPhearson, Samantha Patinella, Krish Ramalingam, Andrea Silverman, Fabrizio Spagnolo, Jasmin Torres, Arvind Varsani, Peter Williamsen for support, advice, discussions, and feedback. This work was funded in part by the New York City Department of Environmental Protection. The Water Research Foundation, the NSF Research Coordination Network for Wastewater Surveillance for SARS-CoV-2 and Qiagen Inc. provided resources, materials and supplies, technical support, and community support.

## Author Contributions

**Conceptualization:** MT, DSS, JJD; **Data Curation:** MT, KC, TG, SK, KMS, DSS, JJD; **Formal Analysis:** MT, IH, SK, DSS, JJD; **Funding Acquisition:** MT, DSS, JJD; **Investigation:** MT, KC, TG, SK, NK, KMS, DSS; **Methodology:** MT, KC, TG, SK, KMS, DSS, JJD; **Project Administration:** MT, DSS, JJD; **Resources:** MT, DSS, JJD; **Supervision:** MT, DSS, JJD; **Visualization:** JJD; **Writing – Original Draft Preparation:** MT, DSS, JJD; **Writing – Review & Editing:** MT, IH, SK, NK, DSS, JJD.

## Supporting Information

S1: Step-by-step protocol, also available on <u>protocols.io</u>

<u>S2: Video showing the resuspension of the pellet with safranin</u>

## Notes

### Competing Interest Statement

We were awarded a next-generation sequencing sponsorship from Qiagen Inc. that included materials and supplies used in isolating RNA. This sponsorship did not influence the results and conclusions reported in this paper.

### Funding Statement

Funding for this project was provided by the New York City Department of Environmental Protection. Qiagen Inc. provided materials and supplies as part of an NGS sponsorship.

### Author Declarations

Queens College Institutional Biosafety Committee approval Protocol IBC-03.

